# Nutrients or processing? An analysis of food and drink items from the UK National Diet and Nutrition Survey based on nutrient content, the NOVA classification, and front of package traffic light labelling

**DOI:** 10.1101/2023.04.24.23289024

**Authors:** Samuel J. Dicken, Rachel L. Batterham, Adrian Brown

## Abstract

**Objective:** To compare food and drink in the UK National Diet and Nutrition Survey (NDNS) database based on their front of package label multiple traffic light (FOPL MTL) score, nutrient content and NOVA classification.

**Design:** National cross-sectional nutrient profiling analysis.

**Setting:** The UK food and drink supply. Food and drink items were obtained from Intake24, the electronic dietary assessment method used in NDNS Year 12 (2019–20).

**Main outcome measures:** Nutrient content, FOPL MTL and the NOVA classification of each item. Items were coded into minimally processed food (MPF), processed culinary ingredients (PCI), processed food (PF) and ultra-processed food (UPF) according to the NOVA classification, and coded into green, amber and red FOPL traffic lights according to Food Standards Agency guidance on fat, saturated fat, total sugar and salt content per 100g.

**Results:** Out of 2,980 items, 55.4% were UPF, 33.1% were MPF, 9.5% were PF, and 2.0% were PCI. UPFs contained greater fat, saturated fat, total sugar, and salt per 100g than MPFs, and had a higher energy density and greater proportion of hyper-palatable items (p<0.001). PFs contained more fat, saturated fat, salt and energy per 100g than MPFs (all p<0.001), but a similar amount of total sugar. UPFs had higher odds of containing red FOPL (odds ratio (OR): 4.59 [95%CI: 3.79 to 5.57]), lower odds of containing green FOPL (OR: 0.05 [95%CI: 0.03, 0.10]), and higher odds of an unhealthier overall FOPL MTL score (OR: 7.0 [95%CI: 6.1 to 8.2], compared with MPFs. When considering items without any red traffic lights, UPF still contained more fat, saturated fat, total sugar and salt than MPFs, and had a higher energy density and greater proportion of hyper-palatable items (p < 0.001). However, a number of UPFs have healthier FOPL MTL scores.

**Conclusions:** Most items in the UK are UPF. UPFs have an unhealthier nutritional profile than MPFs, are more likely to have an unhealthier FOPL MTL score and be more energy-dense and hyper-palatable. When considering items without any red FOPL, UPFs still have a poorer nutritional profile than MPFs, with a higher energy density and hyper-palatability. But, not all UPFs were unhealthy according to FOPL. The results have important implications for understanding how consumers may interpret the healthiness of UPFs or FOPL MTLs, and updating UK food and drink labelling.

What is known:

- Nutrient content is an important determinant of diet-related health. Nutrient content is reflected in UK national dietary guidelines, and at point of purchase through front of package label multiple traffic light (FOPL MTL) scores for nutrients of concern.
- Higher intakes of ultra-processed food (UPF), as defined by the NOVA classification, are associated with higher risks of adverse health outcomes including obesity and cardiometabolic disease.
- The overlap between the nutrient content and FOPL MTLs of the UK food and drink supply with food processing is unknown. How FOPL MTLs might be used to guide consumer purchasing behaviour of UPFs is unclear.

What this study adds:

- This is the first study to compare the nutritional characteristics of food and drink items representative of the UK supply with the NOVA classification.
- There is partial overlap between FOPL MTL and NOVA; UPFs tend to have an unhealthier nutrient profile, but a considerable number of UPFs are considered healthy, based on their FOPL MTL score.
- UPFs also tend to be more energy dense and hyper-palatable than MPFs, even when considering only ‘healthy’ items (without any red FOPL traffic lights).

## Introduction

Diets high in saturated fat, added sugar and salt are associated with increased risks of obesity and non-communicable disease (1–3). As such, UK dietary guidelines recommend that the public reduces their intake of these nutrients, to lower the risk of developing non-communicable disease, including type 2 diabetes, cardiovascular disease and all-cause mortality (4,5).

Dietary advice is communicated to the public through multiple strategies including the Eatwell Guide (EWG), and front of package labelling (FOPL), which is used to help guide consumer choice at the point of purchase (6,7). FOPL systems differ across countries, from simple non-interpretive nutrient information, to interpretive semi-directive colour coded nutrient information (e.g. multiple traffic light (MTL) system in the UK), to interpretive directive advice to support consumer choices (e.g. Nutri-Score in Europe) (8). Current FOPLs focus on the energy and nutrient content of products. Compared with no label, FOPL systems help consumers to better rank the healthiness of food products (9). In the UK, the EWG advice is provided through the semi-directive MTL system, which assigns a green, amber or red colour on a FOPL based on whether the content of fat, saturated fat, salt or sugar is low, medium or high, respectively (6).

Besides nutrient content, mounting evidence shows that processing may also impact on health. The NOVA classification categorises food and drink into four groups: minimally processed food (MPF), processed culinary ingredients (PCI), processed food (PF) and ultra-processed food (UPF) (10). Of particular interest are UPFs, which are industrially reformulated products, typically with five or more ingredients resulting in highly palatable, long lasting, readily accessible, cheap products. UPF now constitutes a significant proportion of adult daily energy intake (11), with nearly 60% of intake in UK adults (12). Such a large intake from UPF is of concern, due to the fact that greater UPF consumption is associated with increased risk of obesity, non-communicable disease and all-cause mortality (13,14).

Indeed, high-UPF diets tend to display poorer nutritional profiles, being higher in fat, saturated fat, free sugar, and lower in fibre, protein and micronutrients (11). However, evidence suggests that the associations between UPF and negative health outcomes appears to be independent of the nutrient content of the diet, or the overall diet pattern (13,14). In the Italian Moli-Sani cohort, both lower dietary nutritional quality (measured as Nutri-score) and greater UPF intake were independently associated with increased risks of cardiovascular and all-cause mortality (15). Further joint analyses showed an attenuated effect of Nutri-Score with mortality, but not of UPF (15). Such findings indicate that dietary nutritional quality and the extent and purpose of processing may capture different, but complimentary components/aspects of diet on health.

A number of potential mechanisms have been suggested to explain the adverse impacts of a high-UPF diet, relating to their typically poorer nutritional quality and aspects of their processing. In particular, UPFs may be more energy dense (16) and have addictive-like, hyperpalatable properties that may encourage increased energy intake (14,17,18), which may be linked to their negative impact on health. The lower protein content of an ultra-processed diet has also been suggested to be a factor driving excess consumption (14,19).

Studies outside of the UK have compared the NOVA classification with nutrient indices such as the Nutrient Rich Food Index (20), FOPL tools such as Nutri-Score or MTL, or nutrient profiling models such as those by the World Health Organization (21–26). Such studies find an inverse, partial association between UPF intake and dietary quality, or typically poorer FOPL profiling in UPFs compared with MPFs, but also find that not all UPFs are nutritionally inferior. No study, however, has examined how the nutrient content of a nationally representative sample of foods and drinks in the UK and their FOPL MTL score varies, based on the NOVA classification. In addition, the overlap between indices of hyper-palatability and the NOVA classification has not been fully determined (18), nor has the energy density profile of UK food and drinks across the NOVA classification This is important to determine whether FOPL MTLs adequately capture aspects of food processing, and whether food processing groups differ nutritionally in the UK. The aim of this study was to examine the association between the extent and purpose of processing of UK food and drinks, their nutrient content, and FOPL MTL scoring. The objective was to compare the NOVA classification of food and drink items from the National Diet and Nutrition Survey (NDNS) Rolling Programme Year 12 database with their FOPL MTL scoring and nutritional characteristics.

## Methods

### Data sources

NDNS is a repeated cross-sectional survey, providing detailed dietary intake from a nationally representative sample of the UK population, aged 1.5 years and older and living in private households, since 2008 (27). Years 1 to 11 (2008/9 to 2018/19) of the survey were assessed using food records across four consecutive days. In Year 12 (2019 to 2020), four-day food records were replaced with four non-consecutive, multiple-pass, 24-hour recalls as the dietary assessment method in the NDNS survey (28). Recalls were conducted using Intake24, a web-based, automated, self-administered 24-hour dietary recall tool (https://intake24.co.uk) (29). Food and drink item names and subgroups used in Intake24 were obtained from the Intake24 team. The corresponding NDNS nutrient databank with the most up-to-date public nutrient information for each food item was obtained from the UK Data Service (https://beta.ukdataservice.ac.uk). Further details on Intake24 and NDNS Year 12 have been published elsewhere (28).

### NOVA classification

Food and drink items were coded according to the NOVA classification (10) (see Supplementary Materials). Years 1 to 11 of NDNS included a nutrient database with mixed dishes separated into their constituent ingredients (e.g. lettuce, mushrooms and mayonnaise from a salad mixed dish), which has been previously coded (30). In NDNS Year 12, the food database was updated by removing redundant items and the nutrient databank was updated to reflect the most current food composition data (28). Food items were no longer disaggregated. For example, the nutrient databank contains sandwiches and salads, rather than the individual components of sandwiches and salads, as in previous NDNS assessments.

Initial coding of the Year 12 dataset was conducted by SD. Each food and drink item was individually coded. Classification was determined based on the NOVA classification definitions (10), item name, subgroup code, best representation from products available in leading UK supermarkets, and the NOVA group of the corresponding item in the NDNS Year 1 to 11 database. The coding, alongside ambiguous food items and food groups were discussed and checked with a Registered Senior Specialist Dietitian (AB), and both authors agreed on the classification. Where it was unspecified or ambiguous as to whether a food item was home-made or ready-made, the authors agreed on the most appropriate classification based on the most likely method of obtaining or preparing the food, by reflecting on the range of, and ingredients within, corresponding products sold from leading UK supermarkets. Further details on the coding of items are provided in the supplementary materials.

### FOPL classification

The NDNS nutrient databank was coded into FOPL MTLs according to Department of Health and Food Standards Agency guidance for fat, saturated fat, total sugar and salt content (6). Items with low content for a given nutrient are coded green, moderate content as amber, and high content as red. The nutrient cut-offs are provided in the Supplementary Materials. To allow for comparability, items were coded per 100g of food or drink. As per FOPL guidance, drink items included lower cut-offs for amber or red colour coding per 100g of drink, which was assumed equivalent to 100ml of drink.

### Statistical analysis

Normally distributed variables were described using means and confidence intervals, and variables that were not normally distributed were described using medians and interquartile ranges (IQR). Categorical variables were described using counts and percentages. Comparisons of non-parametrically distributed nutrient variables between NOVA groups were analysed using Kruskal-Wallis analysis of variance, with Bonferroni correction for multiple comparisons. Categorical variables were analysed using chi-square tests.

The number of products within each NOVA group, the average nutrient and energy content per 100g and distribution of FOPL traffic lights were described. The average nutrient content per 100g and the distribution of individual traffic lights for each nutrient was then compared between NOVA groups. FOPL traffic lights for all four nutrients (fat, saturated fat, total sugar and salt) combined (i.e., MTLs), were then compared. The presence of any red or green FOPL traffic lights (vs. no red or green FOPL traffic lights), the number of red FOPL traffic lights and the number of green FOPL traffic lights (none, one, two, three or four), and the distribution of the overall FOPL MTL score (an 8-level categorical measure ranging from four green to four red FOPL traffic lights (four reds, and three reds and one amber were combined due to few items in these categories)) was then compared across NOVA groups. Due to small numbers of items across categorical levels, PF and PCI were grouped for the ordinal analyses, and for the binary green vs. no green FOPL traffic light analysis.

Unadjusted regression analyses were then conducted to examine the relationship between NOVA groups as the categorical independent variable, and FOPL traffic lights or nutrient content as the dependent variables. Binary regression was used to analyse the odds of containing at least one red traffic light (vs. no red traffic light) and the odds of containing at least one green traffic light (vs. no green traffic light). Ordinal logistic regression was used to model the odds of containing a higher number of red traffic lights, the odds of containing a higher number of green traffic lights, and to model the odds of having a better overall FOPL MTL score.

Subgroup analyses then compared the nutrient and energy content of products with healthier vs. less healthy FOPL MTLs, firstly within UPFs (i.e., healthy vs. unhealthy products), and then between NOVA food groups (i.e., healthy products across NOVA groups). Based on previous evidence suggesting that UK consumers more readily avoid red traffic lights over choosing green traffic lights when choosing healthier products (31,32), items were stratified by the presence or absence of a red FOPL traffic light across the four nutrients. Comparisons between UPFs with or without a red FOPL traffic light were analysed using Mann-Whitney U tests.

To consider the wider nutritional characteristics of food items proposed as potential mechanisms of UPFs in the NDNS database, items were characterised by previously defined quantifiable definitions of hyper-palatable foods (HPF) based on a systematic review of descriptive definitions of hyper-palatability (33). Items were classified into three clusters, containing: (1) fat and sodium (>25% kcal from fat, ≥0.30% sodium content by weight); (2) fat and simple sugars (>20% kcal from fat, >20% kcal from sugar); and (3) carbohydrates and sodium (>40% kcal from carbohydrates, ≥0.20% sodium by weight) (33). The proportions of HPFs were then compared across NOVA groups. Drinks items were excluded from the HPF analysis, as the definition is only applicable to food items.

Items outside of the NOVA classification were removed prior to analysis.

### Sensitivity analysis

The nutrient and energy content of items across NOVA groups were analysed with a binary regression modelling the odds of containing above average nutrient content across the database (i.e., an above median vs. median or below nutrient content. Linear regression was also used to determine the association between NOVA group and the number of red or green FOPL traffic lights. The full FOPL MTL score analysis was repeated using linear regression (where a green FOPL traffic light scored 1, amber scored 2, and red scored 3, for a combined continuous score ranging from 4 (four greens) to 12 (four reds)) and repeated using binary regression to model the odds of an above average (above median vs. median or below) overall FOPL MTL score.

In subgroup analyses, comparisons of nutrient and energy content between subgroups with or without a red FOPL traffic light were repeated with a binary regression, to model the odds of an above average nutrient or energy content (above median vs. median or below value). Subgroup analyses were also repeated by further stratifying products based on the presence of two or more green and no red FOPL traffic lights. Statistical significance was set at a p-value <0.05. Data were analysed in SPSS V29.0.

## Results

The Intake24 dataset contained 3105 items; 109 items were designated outside of the NOVA classification (e.g., fish oil supplements and multivitamins). When aligned with the NDNS nutrient databank, a further 16 did not contain a number corresponding to a respective item in the latest version of the databank, leaving a total of 2,980 items in the final analysis. Over half of the food and drink items were UPFs (n=1650, 55.4%), around a third of the items were MPF (n=986, 33.1%), 9.5% were PF (n=283), and 2.0% (n=61) were PCI.

Table 1 presents the average nutrient and energy content of all items, and within each NOVA group. The median content of fat, saturated fat, total sugar, salt and energy per 100g was 5.1g [IQR: 0.8, 13.5], 1.3g [IQR: 0.2, 4.2], 3.2g [IQR: 1.1, 11.1], 0.3g [IQR: 0.05, 0.86] and 181.0 kcal [IQR: 77.0, 320.0], respectively. MPFs had significantly lower average fat, saturated fat and energy content per 100g than other NOVA groups (all p<0.001) (Figure 1a-e). UPFs contained significantly more fat, saturated fat, total sugar, salt and energy per 100g than MPFs (all p<0.001). UPFs had significantly greater average total sugar content per 100g than other NOVA groups (all p<0.001). PCIs had significantly greater energy content per 100g than other NOVA groups (vs. UPF: p=0.009, vs. MPF or PCI: p<0.001). PFs contained significantly more fat, saturated fat, salt, and energy per 100g than MPFs (all p<0.001), but a similar amount of total sugar (p=0.167). The fat, saturated fat, and salt content of PFs did not differ to UPFs, but the energy density was significantly lower (p<0.001). PCIs tended to have the highest average fat and saturated fat per 100g of all NOVA groups, but was not significantly different from UPFs. Sensitivity analysis with binary regression showed similar findings (Supplementary Table 1). UPFs contained a similar amount of protein as MPFs and PFs, and a similar quantity of fibre as MPFs, but more fibre than PFs (p < 0.001), but this was not meaningfully different. UPFs also had a significantly lower water content than MPFs (75.9g/100g [IQR: 63.0, 87.7] vs. 49.3g/100g [IQR: 16.1, 72.7], p<0.001) (Supplementary Table 2).

**Figure 1.**
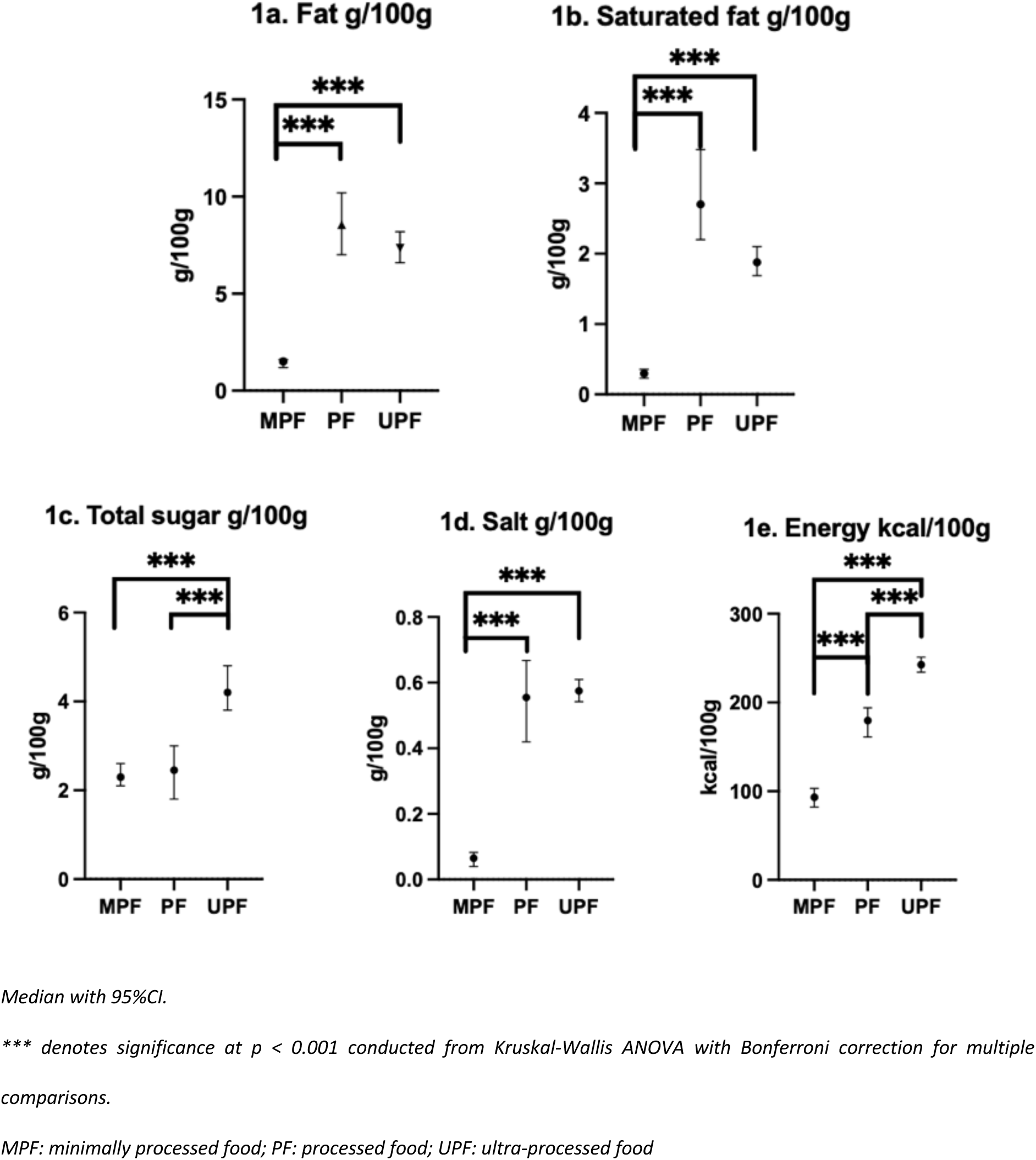
Average fat (1a), saturated fat (1b), total sugar (1c), salt (1d) and energy (1e) content across NOVA food groups (n=2980)

**Table 1:** Fat, saturated fat, total sugar, salt and energy content per 100g by NOVA group.

| Nutrient | Total (n=2,980) |  | MPF (n=986) |  | PCI (n=61) |  | PF (n=283) |  | UPF (n=1650) |  | p-value |
| --- | --- | --- | --- | --- | --- | --- | --- | --- | --- | --- | --- |
|  | Median | IQR | Media<br>n | IQR | Media<br>n | IQR | Media<br>n | IQR | Median | IQR |  |
| Fat (g/100g) | 5.1 | 0.8,<br>13.5 | 1.5 (a) | 0.3, 6.5 | 19.1<br>(b) | 0.0, 98.3 | 8.4 (b) | 0.3,<br>16.4 | 7.4 (b) | 2.1, 15.8 | <0.001 |
| Saturated<br>fat (g/100g) | 1.3 | 0.2,<br>4.2 | 0.3 (a) | 0.07, 1.9 | 12.0<br>(b) | 0.0, 32.5 | 2.7 (b) | 0.1, 5.9 | 1.9 (b) | 0.6, 5.7 | <0.001 |
| Total sugar<br>(g/100g) | 3.2 | 1.1,<br>11.1 | 2.3<br>(ab) | 0.4, 5.1 | 0.0 (a) | 0.0, 2.9 | 2.5 (b) | 0.7, 9.7 | 4.2 (c) | 1.7, 18.9 | <0.001 |
| Salt<br>(g/100g) | 0.27 | 0.05,<br>0.86 | 0.07<br>(a) | 0.01, 0.2 | 0.03<br>(a) | 0.0, 0.12 | 0.56<br>(b) | 0.11,<br>1.05 | 0.58 (b) | 0.18, 1.07 | <0.001 |
| Energy<br>(kcal/100g) | 181.0 | 77,<br>320 | 94.0<br>(a) | 36.0,<br>180.3 | 378.0<br>(b) | 181.0,<br>885.0 | 178.0<br>(c) | 84.0,<br>290.0 | 243.0 (d) | 121.8,<br>371.0 | <0.001 |

Unlike letters indicates significantly different $p < 0.05$
Pairwise comparisons conducted using Kruskal-Wallis ANOVA with Bonferroni correction for multiple comparisons.
IQR: inter-quartile range; MPF: minimally processed food; PCI: processed culinary ingredient; PF: processed food; UPF: ultra-
processed food;

### Traffic light labelling

Items were then coded according to FOPL traffic lights. The total number of red FOPL traffic lights was 18.3% (n=545) for fat, 22.3% (n=665) for saturated fat, 15.7% (n=467) for total sugar and 8.7% (n=259) for salt (Supplementary Table 3). The number of green FOPLs in the database was 40.4% (n=1203) for fat, 50.6% (n=1509) for saturated fat, 59.5% (n=1773) for total sugar and 51.5% (n=1534) for salt. Figure 2a-d presents the percentage of red, amber, and green FOPL traffic lights across fat, saturated fat, total sugar and salt, by NOVA group (PCI not shown). The proportions of green, amber, and red FOPL traffic lights for fat, saturated fat, total sugar, salt significantly differed across NOVA groups (all p<0.001). MPFs had a greater proportion of green FOPL traffic lights and a lower proportion of red FOPL traffic lights, whereas UPFs had a lower proportion of green FOPL traffic light, and a higher proportion of red FOPL traffic light.

**Figure 2.**
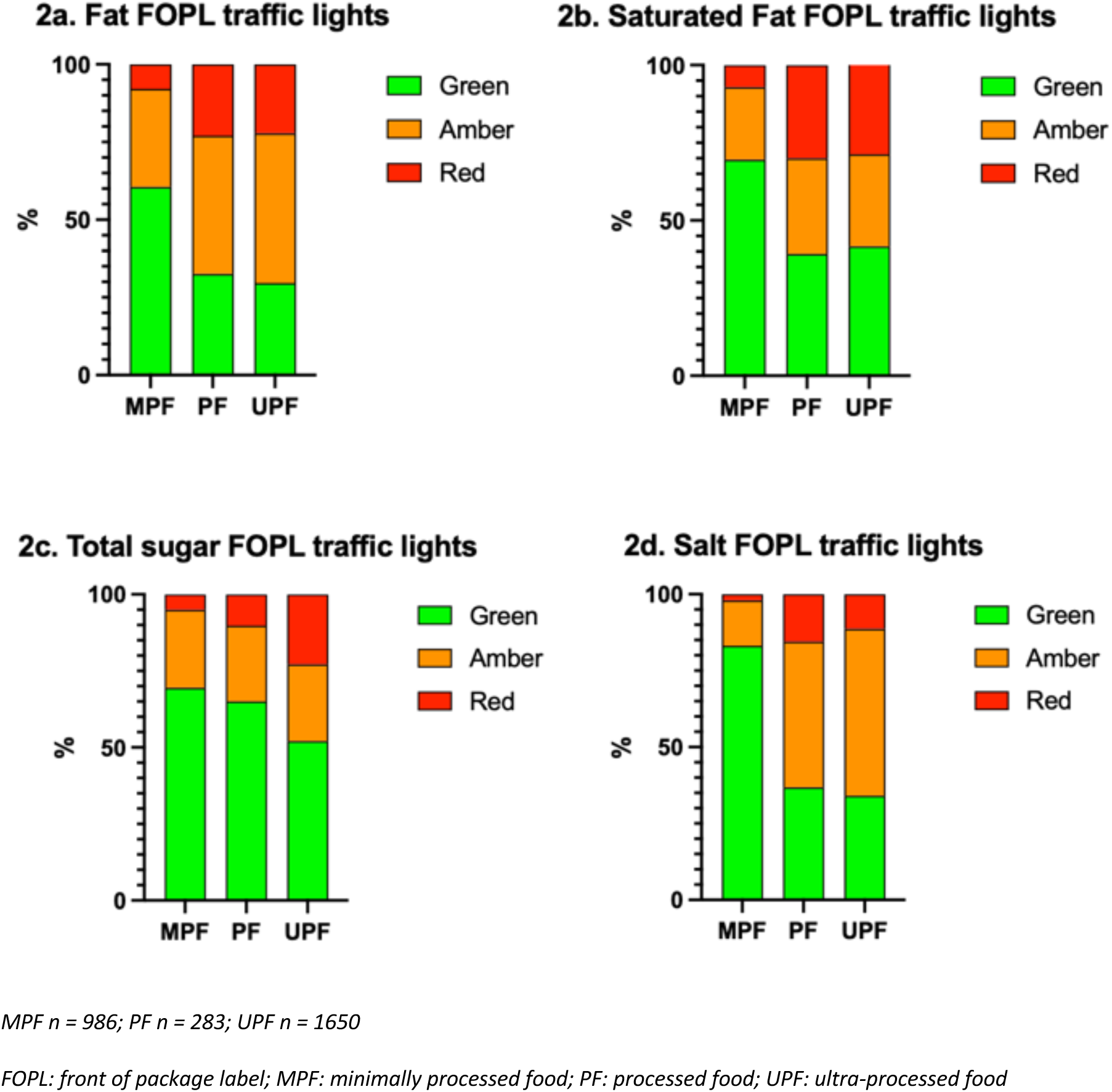
Percentage of red, amber, and green FOPL traffic lights across fat (2a), saturated fat (2b), total sugar (2c) and salt (2d) by NOVA group (PCI not shown).

### Combined FOPL traffic lights

When considering the presence of any red or green FOPL traffic lights per item, approximately two thirds of items contained no red FOPL traffic lights (n=1846, 61.9%), whereas only 9.1% (n=270) of items contained no green FOPL traffic light (Supplementary Table 4). Stratifying by NOVA group, the proportions of items with no red or green FOPL traffic lights significantly differed (p<0.001). Most MPFs contained no red FOPL traffic lights (83.2%; 820 out of 986) with only 16.8% containing one or more red FOPL traffic lights, compared with 51.8% of UPFs containing no red FOPL traffic lights (855 out of 1650), and nearly half with at least one red FOPL traffic light (48.2%; 795 out of 1650). Less than 1% of all MPFs contained no green FOPL traffic light for fat, saturated fat, total sugar, or salt (8 out of 986), compared with 14.0% of UPFs (231 out of 1650). 99.2% of MPFs contained at least one green traffic light, compared with 86.0% of UPFs. In binary regression analyses (Table 2), UPFs had a higher odds of containing one or more red FOPL traffic lights compared with MPFs (OR: 4.59 [95%CI: 3.79, 5.57]), as did PFs (OR: 3.69 [95%CI: 2.77, 4.92] and PCIs (OR: 28.54 [95%CI: 13.80, 59.05]) (p<0.001). Similarly, UPFs had a lower odds of containing one or more green FOPL traffic light compared with MPFs (OR: 0.05 [95%CI: 0.03, 0.10]), as did PCIs and PFs (combined, OR: 0.08 [95%CI: 0.04, 0.18] p<0.001).

**Table 2:** Binary regression modelling the association between NOVA group and the presence of one or more red/green FOPL traffic lights vs no red/green FOPL traffic lights.

|  | Exp(Beta) | 95% Confidence Interval |  | p-value |
| --- | --- | --- | --- | --- |
|  |  | Lower | Upper |  |
| One or more red vs. no red FOPL traffic lights |  |  |  |  |
| UPF | 4.593 | 3.788 | 5.569 | <0.001 |
| PF | 3.690 | 2.765 | 4.924 | <0.001 |
| PCI | 28.541 | 13.795 | 59.047 | <0.001 |

| One or more green vs. no green<br>FOPL traffic lights |  |  |  |  |
| --- | --- | --- | --- | --- |
| UPF | 0.050 | 0.025 | 0.102 | <0.001 |
| PF and PCI | 0.083 | 0.038 | 0.182 | <0.001 |

Reference = MPF
Higher score indicates greater odds of having red or green FOPL traffic lights vs. no red or green FOPL traffic lights
FOPL: front of package label; IQR: inter-quartile range; MPF: minimally processed food; PCI: processed culinary ingredient;
PF: processed food; UPF: ultra-processed food

### Ordinal FOPL MTL score

When considering the number of items with at least one red FOPL traffic light (n=1134, 38.1% of items), 18.6% (n=555) items had one red traffic light, 12.1% (n=360) with two, 7.2% (n=215) with three, and 0.1% (n=4) with four red traffic lights (Supplementary Table 5). When considering the number of items with at least one green traffic light (n=2710, 90.9% of items), 32.9% (n=980) contained one, 19.8% (n=589) contained two, 23.6% (n=703) contained three and 14.7% (n=438) contained four green traffic lights.

Figure 3a-d shows the number and percentage of items with none, one, two, three or four red or green FOPL traffic lights, stratified by NOVA group (PCI not shown) (by MPF and UPF only, in Supplementary Figure 1a-b). The majority of items with one (67.0%, 370 out of 555), two (67.0%, 242 out of 360) or three (84.0%, 181 out of 215) red traffic lights were UPFs. Nearly half of all UPFs (42.8%, 706 out of 1650) contained one green traffic light, followed by two green traffic lights (19.9%, 329 out of 1650), then three (18.4%, 304 out of 1650), then zero green traffic lights (14.0%, 231 out of 1650), with the fewest having four green traffic lights (4.8%, 80 out of 1650). In contrast, most MPFs contained four (33.8%, 333 out of 988), then three (30.9%, 305 out of 986), then two (20.4%, 201 out of 986), and then one green traffic light (14.1%, 139 out of 986).

**Figure 3.**
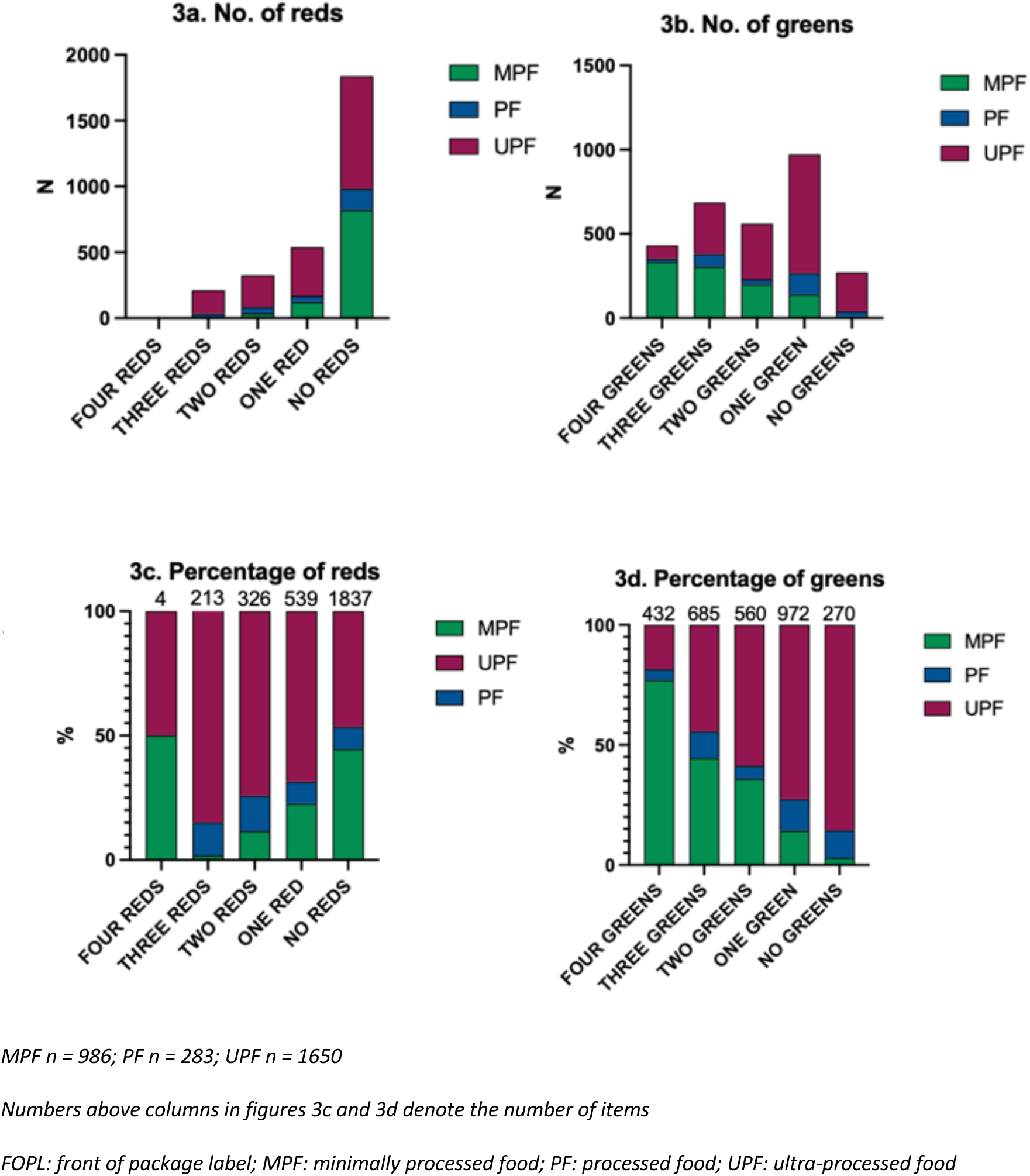
The number (3a and 3b) and percentage (3c and 3d) of total red (3a ad 3c) or green (3b and 3d) FOPL traffic lights for fat, saturated fat, total sugar, and salt, stratified by NOVA group.

The proportions of the number of red or green FOPL traffic lights significantly differed according to NOVA group (both p<0.001) (Supplementary Table 5). UPFs had a higher proportion of one, two or three/four red traffic lights, whereas MPFs had a lower proportion of one, two or three/four red traffic lights. UPFs had a lower proportion of three or four green traffic lights, and a higher proportion of one green or no green traffic lights, whereas MPFs had a higher proportion of three or four green traffic lights, and a lower proportion of one green or no green traffic lights.

Ordinal regression showed that UPFs had a significantly higher odds of containing a greater number of red FOPL traffic lights compared with MPFs (p<0.001, Table 3). UPFs had a 4.84 times [95%CI: 4.00, 5.86] higher odds of containing a greater number of red FOPL traffic lights compared with MPFs. For the number of green FOPL traffic lights, UPFs had a lower odds of containing a greater number of green FOPL traffic lights compared with MPFs. MPFs had 7.30 times [95%CI: 6.25, 8.55] higher odds of containing a greater number of green FOPL traffic lights compared with UPFs. PCIs and PFs (combined) also had a higher odds of containing a greater number of red and a lower number of green traffic lights compared with MPFs (both p<0.001). In sensitivity analyses, results were similar when the number of red or green FOPL traffic lights was modelled as a 5-level continuous score (Supplementary Table 6), where UPFs had 1.25 [95%CI: -1.34, -1.16] fewer green FOPL traffic lights, and 0.63 [95%CI: 0.56, 0.70] more red FOPL traffic lights.

**Table 3.** Ordinal regression modelling the association between NOVA group and the presence of an increasing number of red/green FOPL traffic lights.

|  | Exp(B) | 95% Confidence Interval |  | p-value |
| --- | --- | --- | --- | --- |
|  |  | Lower | Upper |  |
| **Red FOPL traffic lights** |  |  |  |  |
| UPF | 4.842 | 4.002 | 5.857 | <0.001 |
| PF and PCI | 5.461 | 4.212 | 7.079 | <0.001 |
| **Green FOPL traffic lights** |  |  |  |  |
| UPF | 0.137 | 0.117 | 0.160 | <0.001 |
| PF and PCI | 0.212 | 0.169 | 0.266 | <0.001 |

Reference = MPF
Higher score indicates greater odds of having an increasing number of red/green FOPL traffic lights
FOPL: front of package label; IQR: inter-quartile range; MPF: minimally processed food; PCI: processed culinary ingredient;
PF: processed food; UPF: ultra-processed food

### Full FOPL MTL score

As a total FOPL MTL score (ranging from 4 (four green traffic lights) to 12 (four red traffic lights)), the median score was 6.0 [IQR: 5.0, 8.0], corresponding to a FOPL with either two amber and two green traffic lights, or three green traffic lights and one red traffic light (AAGG or GGGR). The profile of FOPL MTLs stratified by NOVA group is presented in Figure 4 (by MPFs and UPFs only, in Supplementary Figure 2). The proportions of FOPL MTLs significantly differed across NOVA groups (p<0.001, Supplementary Table 7). Ordinal regression of the FOPL MTL score showed that UPFs had 7.06 times [95%CI: 6.06, 8.24] higher odds of having an unhealthier FOPL MTL score compared with MPFs (p<0.001, Table 4). PFs and PCIs combined also had a higher odds of an unhealthier FOPL MTL score compared with MPFs (OR: 5.95 [95%CI: 4.74, 7.46]). In sensitivity analyses, results were similar when the FOPL MTL score was modelled as an 8-level continuous outcome ranging from four greens to four reds/three reds and one amber (a score from 4 to 11) (Supplementary Table 8), where UPFs had a FOPL MTL score that was 1.89 [95%CI: 1.75, 2.02] points higher than MPFs (equivalent to nearly two green traffic lights replaced with two amber traffic lights, or one green traffic light replaced with one red traffic light), and when modelled as a binary outcome (above median vs. median and below FOPL MTL score), where UPFs had 6.03 times [5.03, 7.24] higher odds of having an unhealthier FOPL MTL score (Supplementary Table 9).

**Figure 4.**
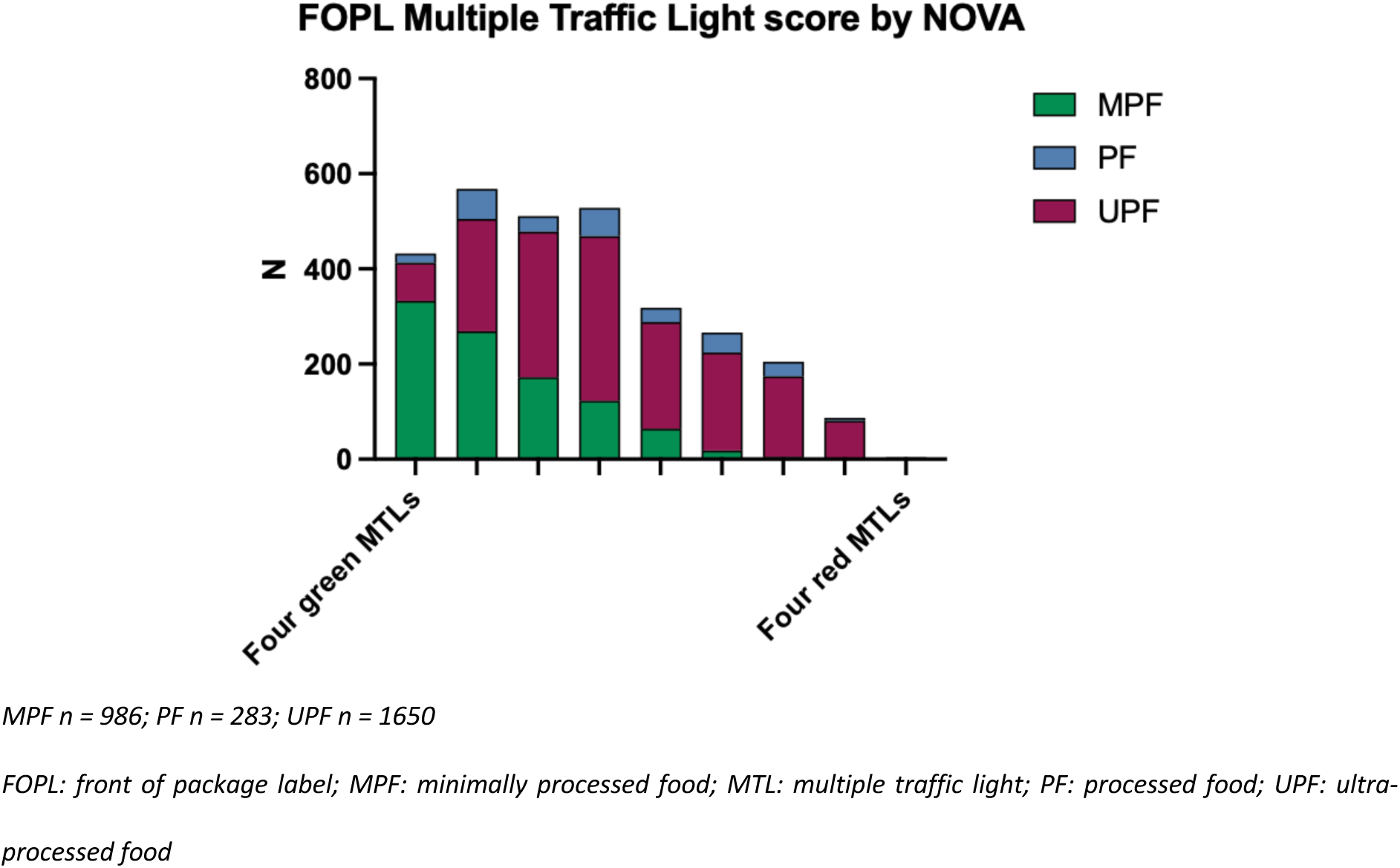
Profile of FOPL MTLs stratified by NOVA group.

**Table 4.** Ordinal regression modelling the association between NOVA group and the presence of an increasing FOPL MTL score.

|  | Exp(B) | 95% Confidence Interval |  | p-value |
| --- | --- | --- | --- | --- |
|  |  | Lower | Upper |  |
| UPF | 7.063 | 6.055 | 8.238 | <0.001 |
| PF and PCI | 5.947 | 4.742 | 7.460 | <0.001 |

*Reference = MPF* *Higher score indicates greater odds of having an unhealthier FOPL MTL score* *FOPL: front of package label; IQR: inter-quartile range; MPF: minimally processed food; MTL: multiple traffic light; PCI:* *processed culinary ingredient; PF: processed food; UPF: ultra-processed food*

### Items with no red FOPL traffic light

Subgroup analyses then considered food and drink items containing no red FOPL traffic lights (n=1846, 61.9%), i.e., ‘healthy’ items. 855 (46.3%) were UPFs, 820 (44.4%) were MPFs, 162 (8.8%) were PFs and 9 (0.5%) were PCIs (Supplementary Table 4). The most common UPFs with no red FOPL traffic lights included sandwiches (n=65, 7.6%), high fibre breakfast cereals (n=43, 5.0%), other milks (e.g., plant-based milk alternatives, milkshakes) (n=38, 4.5%) and white bread (not high fibre, not multiseed) (n=35, 4.1%) (Supplementary Table 10).

UPFs with no red FOPL traffic lights contained lower amounts of fat, saturated fat, total sugar, and salt per 100g than UPFs containing at least one red FOPL traffic light (all p<0.001, Supplementary Table 11). UPFs with no red FOPL traffic lights also had a significantly lower energy density (1.52kcal/g [IQR: 0.77, 2.43] vs. 3.53kcal/g [IQR: 2.51, 4.43], p<0.001). There was no significant difference in protein content (6.4g/100g [IQR: 2.1, 10.7] vs. 5.5g/100g [IQR: •, 9.2], p = 0.761), or fibre content (p = 0.435), between UPFs with or without a red FOPL traffic light.

Compared with MPFs with no red FOPL traffic lights, UPFs with no red FOPL traffic lights contained greater quantities of fat, saturated fat, total sugar, and salt (p<0.001) per 100g (Supplementary Table 12). UPFs had a significantly higher energy density than MPFs (1.52kcal/g [IQR: 0.77, 2.43] vs. 0.75kcal/g [IQR: 0.32, 1.34], p < 0.001) and PFs (1.08kcal/g [IQR: 0.59, 1.75] p<0.001). There was no significant difference in protein (p=0.184) or fibre (p=0.231) content between MPFs and UPFs with no red FOPL traffic lights, but the water content of UPFs was significantly lower than that of MPFs (p<0.001). PFs with no red FOPL traffic lights also contained significantly more saturated fat, total sugar, salt, and energy than MPFs with no red FOPL traffic lights. In sensitivity analyses, binary regressions with median cut-off showed similar associations between NOVA groups and nutrient content (Supplementary Table 13). Sensitivity analyses further considered items with no red FOPL traffic lights and two or more green FOPL traffic lights (n=1403, 47.1%) (corresponding to an item with a median or lower FOPL MTL score with no red FOPL traffic lights). 738 (52.6%) were MPFs, 554 (39.5%) were UPFs, 102 (7.3%) were PFs and 9 (0.6%) were PCIs. UPFs with no reds and at least two greens had lower fat, saturated fat, total sugar and salt content per 100g than UPFs with reds or less than two green FOPL traffic lights (all p<0.001) (Supplementary Table 14). UPFs with no reds and at least two greens contained greater quantities of fat, saturated fat, total sugar, and salt than MPFs with no reds and at least two green FOPL traffic lights (p<0.001), and had a significantly higher energy density (1.07kcal/g [IQR: 0.48, 2.21] vs. 0.71kcal/g [IQR: 0.29, 1.24], p<0.001) and PFs (0.74 kcal/g [IQR: 0.40, 1.02], p<0.001) (Supplementary Table 15).

### Hyper-palatable food

When stratified by hyper-palatability, 46.8% (n=1246) of all food items (n=2665) were classified as being hyper-palatable based on their fat, sodium, sugar and carbohydrate content. Of which, 79.8% were UPFs (994 out of 1246). Across each cluster, the majority of HPFs defined by: (1) fat and sodium (78.5%, 504 out of 642), (2) fat and simple sugars (75.5%, 318 out of 421), or (3) carbohydrates and sodium (93.7%, 342 out of 365), were UPFs (Figure 5). Across all items, a significantly greater proportion of UPFs than MPFs were classed as HPF for: (1) fat and sodium, (2) fat and simple sugars, or (3) carbohydrates and sodium, and overall (combining all clusters) (all p <0.001) (Supplementary Table 16). When considering ‘healthy’ food items (i.e. with no red FOPL traffic lights, n= 1620), there was still a significantly greater proportion of UPFs than MPFs that were defined as HPF, based on: (1) fat and sodium, and (3) carbohydrates and sodium, and overall (all p<0.001), but there was no significant difference for (2) fat and simple sugars (p=0.367).

**Figure 5.**
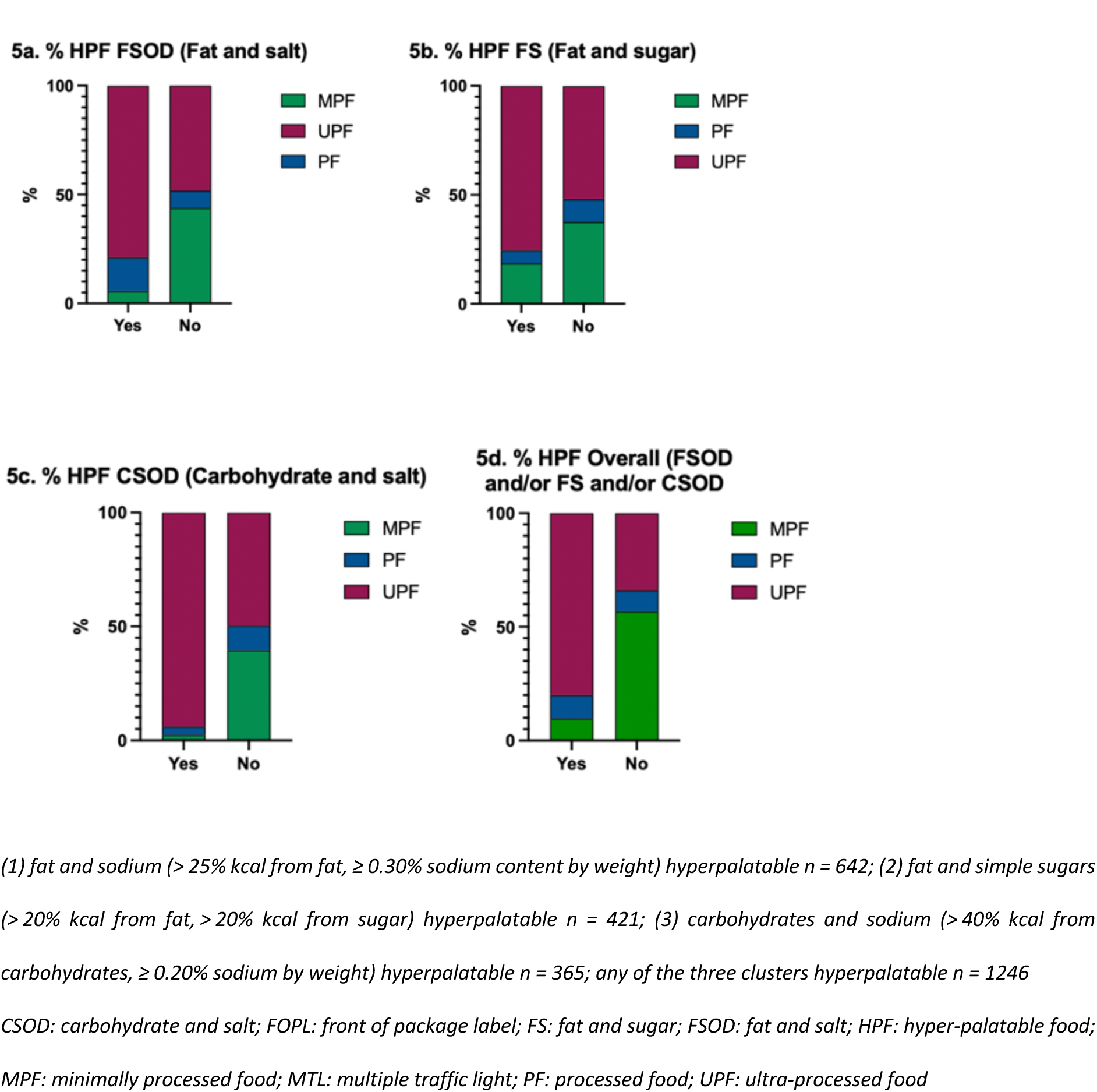
Proportions of food items meeting criteria for hyper-palatability based on: (1) fat and sodium (5a) (2) fat and simple sugars (5b) (3) carbohydrates and sodium (or any of the three clusters (5d) (PCI not shown)

## Discussion

The findings from this analysis indicate that UPFs tend to have an unhealthier nutritional profile than MPFs, but not PFs. UPFs contained greater amounts of fat, saturated fat, total sugar, and salt than MPFs, were more energy dense, and were more likely to be classed as hyper-palatable. Compared with PFs, UPFs contained similar amounts of fat, saturated fat and salt, but greater amounts of sugar, and were more energy dense. UPFs were more likely to have fewer green FOPL traffic lights, a greater number of red FOPL traffic lights, and be rated as unhealthier based on their overall FOPL MTL score. However, not all UPFs had an unhealthy nutrient profile. Over half of UPFs had no red FOPL traffic lights, and a significant number of UPFs had a FOPL MTL score similar to MPFs, with nearly half of items with no red FOPL traffic lights being classed as UPFs. However, UPFs with no red FOPL traffic lights still had a worse nutritional profile and higher energy density than MPFs with an equivalent FOPL MTL score, and were still more likely to be classed as hyper-palatable. These results suggest that the FOPL MTL system does not fully differentiate between UPFs and MPFs, only partially capturing the extent and purpose of food processing.

Aspects of ultra-processing, such as changes to the food matrix, greater energy density and the combination of nutrients not usually found in MPFs have been suggested to alter oro-sensory exposure time, increasing eating rates and resulting in overconsumption (16,18,34). In this study, UPFs were more energy dense than MPFs and PFs. Even when considering only ‘healthy’ items with no red FOPL traffic lights, UPFs still had double the energy density of MPFs (1.52kcal/g vs. 0.75kcal/g). Lowering energy density can lower daily energy intake in a strong and linear fashion (36), with an average 223kcal reduction in energy intake when lowering meal energy density from 1.5 kcal/g to 1.1 kcal/g (37). In a metabolic ward crossover study comparing a 2-week UPF diet (1.36 kcal/g) versus a 2-week MPF diet (1.09 kcal/g) matched for presented energy and macronutrients, participants consumed ∼500kcal per day more on the UPF than MPF diet, resulting in 0.9kg weight gain on the UPF diet, but 0.9kg of weight loss on the MPF diet (14). Hyper-palatability has also been suggested to be important for food choice and consumption (33). Many UPFs have hyper-palatable properties, which have been proposed to have addictive-like qualities by inducing a greater hedonic response when consumed, increasing reward-driven eating (18,33,38). In the metabolic ward study, both energy density (45.1 ± 13.6%) and hyper-palatability (41.9 ± 6.5%) explained large proportions of the greater daily non-beverage energy intake on the UPF versus MPF diet (35). In addition, UPFs have also been described by their extensive matrix degradation, which can make them softer and easier to consume at a faster rate (34). Therefore, UPFs may capture several characteristics that may predispose to overconsumption that are not sufficiently reflected in current FOPL guidance. These findings suggest that within the UK food and drink supply, choosing healthier UPF items based on the FOPL MTL score may still predispose to increased energy intake, compared with MPFs with a similar FOPL MTL score.

The NOVA classification does not explicitly differentiate food and drink based on their nutrient content. But, high-UPF diets tend to be nutritionally poorer (11), evident by the partial overlap with the FOPL MTL score. However, although UPFs tend to have an unhealthier nutritional profile than MPFs, not all do. This finding is in line with studies from other countries using different nutrient indices (21,26). Given the adverse impacts of high intakes of nutritionally poor foods high in fat, sugar and salt (39–41), transnational corporations have reformulated their products to contain less fat, saturated fat, added sugar or salt (42). Previous analyses based on nutrient profiling have suggested that UPFs with better nutritional profiles may be considered healthy (43), particularly if they carry a nutrient or health claim. Notably, a large proportion of UPFs in this study contained one green FOPL traffic light, such as low-fat ready meals, sauces and puddings, which could carry nutrient claims. However, to what extent a diet high in UPF but consisting of items with better nutritional quality and containing nutrition/health claims can constitute a healthy diet remains to be seen.

Whether the inclusion of the NOVA classification and avoidance of UPFs should be recommended in dietary guidelines is currently debated (44). However, the most up-to-date scientific guidance from the American Heart Association and American Society for Preventive Cardiology includes advice to choose MPF and/or minimise UPF intake, alongside standard dietary guidance to limit foods high in fat, sugar and salt (2,3). Such recommendations indicate that consumers should consider not just the nutrient content, but also the processing of their food purchases. FOPLs encourage healthier in-supermarket food purchases (45), and previous studies suggest Nutri-Score to be most effective in improving consumer understanding of the healthfulness of food products in the UK and in Europe (9,46,47). Current FOPLs in the UK use reductive approaches to provide consumers with guidance regarding which products to consume more of, and those to consume less of. FOPLs do not take into account the extent and purpose of food processing, and may be insufficient to help inform consumers to choose MPFs over UPFs. In addition, PFs and UPFs had similar nutrient content and would be expected to carry similar FOPL MTLs. But, UPFs were still more energy dense, potentially due to the significantly lower water content of UPFs. It is therefore important to consider whether current FOPLs provide adequate information to guide consumers towards making healthy in-store food choices. However, the results here also highlight that FOPL MTL scores carry important information not captured by the NOVA classification. Exclusively using the NOVA classification to choose food and drinks might lack the granularity to identify the nutrient quality of items, excluding potentially healthy food choices unnecessarily. Therefore, a combined approach to dietary guidelines and food labelling that considers both the extent and purpose processing and existing indicators of dietary quality may provide the most informative guidance (48). How best to communicate this combined guidance needs to be deliberated, to avoid presenting a complex public health message. Policy makers must also consider the financial implications and abilities of the UK public to shift towards an MPF diet, and away from UPFs particularly with the current cost of living crisis.

## Strengths and limitations

There a several strengths of this study, including the use of a large UK database of nationally representative food and drink items, with a matching nutrient database using average nutrient compositions. Products were compared not only on their nutrient content, but also their FOPL coding used at consumer point of purchase and wider characteristics that influence food intake. Limitations include the analysis per 100g, which although allows for comparability between food and drink items, does not reflect nutrient intakes of actual portions that may be consumed. Other limitations include the categorisation of items into NOVA groups; despite agreement amongst authors on classification, a small number of items could have been entered into more than one group. It was also not possible to assess other properties of items that may influence eating rate and energy intake in this analysis, such as textural properties (34).

## Conclusion

The NOVA classification partially overlaps with the nutrient content of food and drink items available in the UK. UPFs tend to have an unhealthier nutritional profile according to FOPL and a higher energy density compared to MPFs, with a greater number of red FOPL traffic lights, and fewer green FOPL traffic lights. UPFs tended to be nutritionally similar to PFs, but were still more energy dense. However, many UPFs contain no red FOPL traffic lights and multiple green FOPL traffic lights, which have an improved nutritional profile equivalent to many MPFs. However, ‘healthy’ UPFs still tend to contain more fat, saturated fat, total sugar and salt than MPFs with an equivalent FOPL traffic light score, are more energy dense and more likely to be hyper-palatable. These results have important implications for understanding how consumers may interpret the relative healthiness of MPFs and UPFs, and for updating UK food and drink labelling.

## Data Availability

Data can be made available upon request to the authors. For access to Intake24 datasets, please contact the Intake24 team directly; https://intake24.co.uk.

## Acknowledgements

The authors would like to thank the Intake24 team for their administrative support in obtaining the relevant datasets, Fernanda Rauber and her team for sharing their full NOVA classification of food and drink items from NDNS rolling programme Years 1-11, and Helen Croker for providing insightful comments on a draft of the manuscript.

## Patient and Public Involvement

Patients and the public were not involved in the design, conduct, reporting or dissemination of the analysis.

## Author Contributions

S.J.D., data collection and processing, design, data analysis, writing—first draft; R.L.B., writing—editing and reviewing, supervision; A.B. conceptualisation, data processing, writing—editing and reviewing, supervision. All authors have read and agreed to the final manuscript version.

