## Supplementary Materials for "Nutrients or processing? An analysis of food and drink items from the UK National Diet and Nutrition Survey based on nutrient content, the NOVA classification, and front of package traffic light labelling"

**Comparing the nature, extent and purpose of food processing determined by the NOVA classification, and front of package traffic light labelling from the United Kingdom National Diet and Nutrition Survey**

**Supplementary materials**

**NOVA classification (adapted from Monteiro et al., 2019 (1)).**

**Minimally processed foods**

Unprocessed foods altered by industrial processes such as removal of inedible or unwanted parts, drying, crushing, grinding, fractioning, roasting, boiling, pasteurisation, refrigeration, freezing, placing in containers, vacuum packaging, non-alcoholic fermentation, and other methods that do not add salt, sugar, oils or fats or other food substances to the original food.

Examples include fresh, squeezed, chilled, frozen, or dried fruits and vegetables; grains; legumes; meat, poultry, fish; eggs; milk; fruit or vegetable juices (with no added sugar, sweeteners or flavours); flakes or flour made from corn, wheat, oats, or cassava; seeds (with no added salt or sugar); herbs and spices, plain yoghurt; tea, coffee, and drinking water.

**Processed culinary ingredients**

Substances obtained directly from minimally processed foods or from nature by industrial processes such as pressing, centrifuging, refining, extracting or mining. They are used in preparing, seasoning and cooking minimally processed foods.

Examples include vegetable oils; butter and lard; sugar and molasses; honey extracted; starches extracted from corn and other plants, and salt.

**Processed food**

Products made by adding salt, oil, sugar or other processed culinary ingredients to minimally processed foods, using preservation methods such as canning and bottling, or for breads and cheeses, using non-alcoholic fermentation. Processes and ingredients are used to increase the durability of minimally processed foods and make them more enjoyable, by modifying or enhancing sensory qualities.

Examples include canned or bottled vegetables and legumes in brine; salted or sugared nuts and seeds; salted, dried, cured, or smoked meats and fish; canned fish; fruits in syrup and freshly made unpackaged breads and cheeses.

**Ultra-processed food**

Formulations of ingredients mostly of exclusive industrial use, that result from a series of industrial processes. Many processes require sophisticated equipment and technology. Processes enabling the manufacture of ultra-processed foods include fractioning whole foods into substances, chemical modifications of substances, assembly of unmodified and modified food substances using industrial techniques such as extrusion, moulding and pre-frying, frequent application of additives whose function is to make the final product palatable or hyper-palatable (‘cosmetic additives’), and sophisticated packaging, usually with synthetic materials. Ingredients often include sugar, oils and fats, and salt, generally in combination; substances that are sources of energy and nutrients but of no or rare culinary use such as high fructose corn syrup, hydrogenated or interesterified oils, and protein isolates; cosmetic additives such as flavours, flavour enhancers, colours, emulsifiers, sweeteners, thickeners, and anti-foaming, bulking, carbonating, foaming, gelling, and glazing agents; and additives that prolong product duration, protect original properties or prevent proliferation of microorganisms. Processes and ingredients used to manufacture ultra-processed foods are designed to create highly profitable (low cost and long shelf-life), convenient, hyper-palatable snacked products liable to displace all other NOVA food groups, notably minimally processed foods.

Examples include carbonated soft drinks; sweet or savoury packaged snacks; chocolate, confectionery; ice-cream; mass-produced packaged breads and buns; margarines and other spreads; biscuits, pastries, cakes, and cake mixes; breakfast ‘cereals’, ‘cereal’ and ‘energy’ bars; ‘energy’ drinks; milk drinks, ‘fruit’ yoghurts and ‘fruit’ drinks; ‘cocoa’ drinks; ‘instant’ sauces; infant formulas, follow-on milks, other baby products; and ‘health’ and ‘slimming’ products such as meal replacement shakes and powders. Many ready to heat products including pre-prepared pies and pasta and pizza dishes; poultry and fish ‘nuggets’ and ‘sticks’, sausages, burgers, hot dogs, and other reconstituted meat products, and powdered and packaged ‘instant’ soups, noodles and desserts are ultra-processed foods.

**NOVA coding in detail**

Classification was determined by considering the food or drink item name, subgroup code, best representation from leading UK supermarkets, and NOVA code of the corresponding food item in the NDNS Year 1 to 11 database.

- If the item was designated a ‘homemade’ subgroup with no added detail and most likely homemade, it is classed as MPF
- Mixed dishes such as curry, pasta etc if defined as homemade and no other description, they are coded as MPF. If there is a specified PCI, PF or UPF in the food or drink item name, then they are coded as PF or UPF, e.g. homemade cream-based sauces or white wine sauces in a mixed dish, curry in a specified cream/coconut based sauce, or pasta with vegetables in a cream/cheese-based sauce are PF.
- If there is a sauce which is an unspecified recipe and the item is homemade then it is coded as PF, if the sauce is UPF then the dish is UPF.
- If a PF or UPF is fundamental to the dish, it is coded as such - dishes with cheese-based toppings such as lasagne are classed as PF.
- Homemade dishes with mayonnaise (e.g. tuna mayonnaise or potato salad) are PF as a minimum, or UPF. Salads with no dressing are coded as MPF. Unspecified homemade salads or readymade salads with mayonnaise, salad cream or French-dressing are coded as UPF.
- Homemade buns, cakes, pastries, puddings and pancakes are coded as PF, given that they are combinations of MPFs and PCIs. Pies, dumplings or pastry doughs are also PF.
- Homemade stew or meat in gravy (assuming the gravy is from cooking the meat, i.e. use of no OXO cube or gravy granules) is coded as MPF.
- Homemade cottage pie or shepherd’s pie are coded as MPF.
- Homemade battered/breaded fish are coded as PF. Readymade battered/breaded fish are UPF. Unspecified battered/breaded fish are UPF.
- Canned vegetables in an unspecified medium (not described in the food item name or subgroup description) are assumed to be in water and coded as MPF. Stuffed vegetables with an undefined filling are MPF. Canned fruit are assumed to be in a syrup or fruit juice, and therefore are coded as PF.
- Bacon, ham, gammon and similar meats such as deli/pre-packed sliced meat are coded as UPF. Traditional hams (e.g. prosciutto, parma or serrano ham) coded as PF.
- Nut butters are PF.
- Tinned fish in oil, brine or tomato are coded as PF.
- Gluten-free foods are coded as UPF.
- Jams/conserves, marmalades and lemon curd are coded as UPF as typically containing gelling agents in UK supermarkets.
- Breakfast cereals including muesli are coded as UPF. Plain porridge is coded as MPF.
- Items made with a plant-based milk, e.g. lattes/cappuccinos made with plant-based milk or porridge made with plant-based milk are coded as MPF.
- Single cream, double cream and crème fraiche are coded as PCI.
- Chow Mein and Chop Suey (which require soy sauce) are coded as PF. Stir fry (assuming only MPF ingredients) is MPF.

**Front of package label (FOPL) multiple traffic light (MTL) coding**

Coding of FOPL MTLs was conducted according to Department of Health and Food Standards Agency guidance for fat, saturated fat, total sugar and salt content (1) (drinks were coded per 100g instead of per 100ml):

Fat /100g:

Food

- Red: > 17.5g
- Amber: > 3.0g to ≤ 17.5g
- Green: ≤ 3.0g

Drink

- Red: > 8.75g
- Amber: > 1.5g to ≤ 8.75g
- Green: 1.5g

Saturated fat /100g:

Food

- Red: > 5.0g
- Amber: > 1.5g to ≤ 5.0g
- Green: ≤ 1.5g

Drink

- Red: > 2.5g
- Amber: > 0.75g to ≤ 2.5g
- Green: ≤ 0.75g

Total sugar /100g:

Food

- Red: > 22.5g
- Amber: > 5.0g to ≤ 22.5g
- Green: ≤ 5.0g

Drink

- Red: > 11.25g
- Amber > 2.5g to ≤ 11.25g
- Green: ≤ 2.5g

Salt /100g:

Food

- Red: >1.5g
- Amber: > 0.3g to ≤ 1.5g
- Green: ≤ 0.3g

Drink

- Red: > 0.75g
- Amber: >0.3g to ≤0.75g
- Green: ≤ 0.3g

1. Monteiro CA, Cannon G, Levy RB, Moubarac JC, Louzada ML, Rauber F, et al. Ultra-processed foods: what they are and how to identify them. Public Health Nutr. 2019 Apr;22(5):936–41.
2. GOV.UK. Front of Pack nutrition labelling guidance. GOV.UK. [cited 2022 Nov 21]. Available from: https://www.gov.uk/government/publications/front-of-pack-nutrition-labelling-guidance

**Supplementary Tables**

1. **Supplementary Table 1: Binary regression modelling the association between NOVA group and nutrient content (median and below vs. above median for each nutrient)**
2. **Supplementary Table 2: Protein, fibre and water per 100g by NOVA group**
3. **Supplementary Table 3: Fat, saturated fat, total sugar and salt FOPL traffic lights by NOVA group**
4. **Supplementary Table 4: Proportion of items with no red or green FOPL traffic lights by NOVA group**
5. **Supplementary Table 5: Total number of red or green FOPL traffic lights by NOVA group**
6. **Supplementary Table 6: Linear regression modelling the association between NOVA group and the number of green/red FOPL traffic lights**
7. **Supplementary Table 7: Number of items with a given FOPL MTL score by NOVA group**
8. **Supplementary Table 8: Linear regression modelling the association between NOVA group and FOPL MTL score**
9. **Supplementary Table 9: Binary regression modelling the association between NOVA group and FOPL MTL score (median and below vs. above median cutoff)**
10. **Supplementary Table 10: The most common MPFs and UPFs with no red FOPL traffic lights**
11. **Supplementary Table 11: Nutrient content per 100g of UPFs with or without red FOPL traffic lights**
12. **Supplementary Table 12: Nutrient and energy content of items with no red FOPL traffic lights by NOVA group.**
13. **Supplementary Table 13: Binary regression modelling the association between NOVA group and nutrient content of items with no red FOPL traffic lights (median and below vs. above median for each nutrient)**
14. **Supplementary Table 14: Nutrient content per 100g of UPFs without red FOPL traffic lights and two or more green FOPL traffic lights vs. all others**
15. **Supplementary Table 15: Nutrient content per 100g of items without red FOPL traffic lights and two or more green FOPL traffic lights vs. all others, by NOVA group**
16. **Supplementary Table 16: Number of items defined as being hyper-palatable based on fat, sodium, sugar and carbohydrate content.**

**Supplementary Figures**

1. **Supplementary Figure 1: The number of red or green FOPL traffic lights within MPFs or UPFs.**
2. **Supplementary Figure 2: FOPL MTL score within MPFs or UPFs.**

**Supplementary Table 1: Binary regression modelling the association between NOVA group and nutrient content (median and below vs. above median for each nutrient)**

| **Nutrient** | **Exp(B)** | **95% Confidence Interval** | | **p-value** |
| --- | --- | --- | --- | --- |
|  |  | **Lower** | **Upper** |  |
| **Fat** |  |  |  |  |
| UPF | 3.352 | 2.835 | 3.964 | <0.001 |
| PF | 3.779 | 2.868 | 4.980 | <0.001 |
| PCI | 3.309 | 1.951 | 5.610 | <0.001 |
| **Saturated Fat** |  |  |  |  |
| UPF | 3.243 | 2.743 | 3.835 | <0.001 |
| PF | 3.892 | 2.952 | 5.133 | <0.001 |
| PCI | 3.357 | 1.980 | 5.692 | <0.001 |
| **Total Sugar** |  |  |  |  |
| UPF | 1.931 | 1.646 | 2.267 | <0.001 |
| PF | 1.043 | 0.798 | 1.364 | 0.757 |
| PCI | 0.435 | 0.236 | 0.800 | 0.007 |
| **Salt** |  |  |  |  |
| UPF | 9.589 | 7.887 | 11.659 | <0.001 |
| PF | 8.502 | 6.339 | 11.404 | <0.001 |
| PCI | 1.210 | 0.630 | 2.324 | 0.568 |
| **Energy** |  |  |  |  |
| UPF | 5.449 | 4.569 | 6.499 | <0.001 |
| PF | 2.935 | 2.231 | 3.862 | <0.001 |
| PCI | 9.326 | 5.116 | 17.000 | <0.001 |

Reference = MPF

Higher score indicates greater odds of having above median nutrient content vs. median or below

**Supplementary Table 2: Protein, fibre and water per 100g by NOVA group**

| **Nutrient** | **TOTAL** | | **MPF** | | **PCI** | | **PF** | | **UPF** | |  |
| --- | --- | --- | --- | --- | --- | --- | --- | --- | --- | --- | --- |
|  | **Median** | **IQR** | **Median** | **IQR** | **Median** | **IQR** | **Median** | **IQR** | **Median** | **IQR** | **p-value** |
| **Protein (g/100g)** | 5.3 | 1.7, 11.1 | 3.5 (a) | 1.4, 14.0 | 0.3 (b) | 0.0, 1.8 | 7.4 (c) | 1.2, 15.3 | 5.9 (ac) | 2.6, 10.2 | <0.001 |
| **Fibre (g/100g)** | 1.4 | 0.1, 2.8 | 1.6 (a) | 0.0, 3.0 | 0.0 (b) | 0.0, 0.0 | 1.0 (c) | 0.0, 1.6 | 1.5 (a) | 0.5, 2.9 | <0.001 |
| **Water (g/100g)** | 62.1 | 28.7, 79.9 | 75.9 (a) | 63.0, 87.7 | 5.0 (b) | 0.0, 32.1 | 65.6 (c) | 45.7, 79.0 | 49.3 (d) | 16.1, 72.7 | <0.001 |

Unlike letters indicates significantly different p < 0.05

Pairwise comparisons conducted using Kruskal-Wallis ANOVA with Bonferroni correction for multiple comparisons.

**Supplementary Table 3: Fat, saturated fat, total sugar and salt FOPL traffic lights by NOVA group**

|  | **MPF** | **PCI** | **PF** | **UPF** | **Total** | | **p-value** |
| --- | --- | --- | --- | --- | --- | --- | --- |
| **Fat** |  |  |  |  | **n** | **%** |  |
| Green | 597 | 25 | 92 | 489 | 1203 | 40.4 |  |
| Amber | 311 | 0 | 126 | 795 | 1232 | 41.3 |  |
| Red | 78 | 36 | 65 | 366 | 545 | 18.3 | < 0.001 |
| **Saturated fat** |  |  |  |  |  |  |  |
| Green | 686 | 25 | 111 | 687 | 1509 | 50.6 |  |
| Amber | 230 | 0 | 87 | 489 | 806 | 27.0 |  |
| Red | 70 | 36 | 85 | 474 | 665 | 22.3 | < 0.001 |
| **Total sugar** |  |  |  |  |  |  |  |
| Green | 685 | 45 | 184 | 859 | 1773 | 59.5 |  |
| Amber | 251 | 5 | 70 | 414 | 740 | 24.8 |  |
| Red | 50 | 11 | 29 | 377 | 467 | 15.7 | < 0.001 |
| **Salt** |  |  |  |  |  |  |  |
| Green | 820 | 49 | 104 | 561 | 1534 | 51.5 |  |
| Amber | 146 | 5 | 135 | 901 | 1187 | 39.8 |  |
| Red | 20 | 7 | 44 | 188 | 259 | 8.7 | < 0.001 |
|  | 986 | 61 | 283 | 1650 | 2980 |  |  |

**Supplementary Table 4: Proportion of items with no red or green FOPL traffic lights by NOVA group**

|  | **MPF** | **PCI** | **PF** | **UPF** | **p-value** | **Total** | |
| --- | --- | --- | --- | --- | --- | --- | --- |
|  |  |  |  |  |  | **n** | **%** |
| **No red FOPL traffic lights** | 820 | 9 | 162 | 855 |  | 1846 | 61.9 |
| **One or more red FOPL traffic lights** | 166 | 52 | 121 | 795 | < 0.001 | 1134 | 38.1 |
| **No green FOPL traffic lights** | 8 | 0 | 31 | 231 |  | 270 | 9.1 |
| **One or more green FOPL traffic lights** | 978 | 61 | 252 | 1419 | < 0.001 | 2710 | 90.9 |
|  | 986 | 61 | 283 | 1650 |  | 2980 |  |

**Supplementary Table 5: Total number of red or green FOPL traffic lights by NOVA group**

|  | **MPF** | **PCI** | **PF** | **UPF** | **Total** | | **p-value** |
| --- | --- | --- | --- | --- | --- | --- | --- |
|  |  |  |  |  | **n** | **%** |  |
| FOUR REDS | 2 | 0 | 0 | 2 | 4 | 0.1 |  |
| THREE REDS | 4 | 2 | 28 | 181 | 215 | 7.2 |  |
| TWO REDS | 38 | 34 | 46 | 242 | 360 | 12.1 |  |
| ONE RED | 122 | 16 | 47 | 370 | 555 | 18.6 |  |
| NO REDS | 820 | 9 | 162 | 855 | 1846 | 61.9 | <0.001 |
| FOUR GREENS | 333 | 6 | 19 | 80 | 438 | 14.7 |  |
| THREE GREENS | 305 | 18 | 76 | 304 | 703 | 23.6 |  |
| TWO GREENS | 201 | 29 | 30 | 329 | 589 | 19.8 |  |
| ONE GREEN | 139 | 8 | 127 | 706 | 980 | 32.9 |  |
| NO GREENS | 8 | 0 | 31 | 231 | 270 | 9.1 | <0.001 |
|  | 986 | 61 | 283 | 1650 | 2980 |  |  |

**Supplementary Table 6: Linear regression modelling the association between NOVA group and the number of green/red FOPL traffic lights**

|  | **Beta** | **95% Confidence Interval** | | **p-value** |
| --- | --- | --- | --- | --- |
|  |  | **Lower** | **Upper** |  |
| **Number of red FOPL traffic lights** |  |  |  |  |
| UPF | 0.630 | 0.559 | 0.702 | <0.001 |
| PF and PCI | 0.689 | 0.577 | 0.801 | <0.001 |
| **Number of green FOPL traffic lights** |  |  |  |  |
| UPF | -1.254 | -1.340 | -1.168 | <0.001 |
| PF and PCI | -0.982 | -1.115 | -0.848 | <0.001 |

Reference = MPF

Higher score indicates increasing number of red/green FOPL traffic lights

**Supplementary Table 7: Number of items with a given FOPL MTL score by NOVA group**

|  | **MPF** | **PCI and PF** | **UPF** | **Total** | **p-value** |
| --- | --- | --- | --- | --- | --- |
| GGGG | 333 | 25 | 80 | 438 |  |
| AGGG | 269 | 66 | 236 | 571 |  |
| AAGG/GGGR | 172 | 48 | 306 | 526 |  |
| AAAG/GGAR | 123 | 60 | 346 | 529 |  |
| AAAA/RAAG/RRGG | 64 | 58 | 224 | 346 |  |
| RAAA/RRAG | 18 | 48 | 206 | 272 |  |
| RRAA/RRRG | 5 | 33 | 169 | 207 |  |
| RRRA/RRRR | 2 | 6 | 83 | 91 | < 0.001 |
|  | 986 | 344 | 1650 | 2980 |  |

G, Green FOPL traffic light; A, Amber FOPL traffic light; R, Red FOPL traffic light, e.g. GGGG = four green FOPL traffic lights

**Supplementary Table 8: Linear regression modelling the association between NOVA group and FOPL MTL score**

|  | **Beta** | **95% Confidence Interval** | | **p-value** |
| --- | --- | --- | --- | --- |
|  |  | **Lower** | **Upper** |  |
| **FOPL MTL score** |  |  |  |  |
| UPF | 1.885 | 1.750 | 2.021 | <0.001 |
| PF and PCI | 1.672 | 1.461 | 1.884 | <0.001 |

Green FOPL traffic light = 1, Amber FOPL traffic light = 2, Red FOPL traffic light = 3. Four green FOPL traffic lights = 4, four red FOPL traffic lights = 12.

Reference = MPF

Higher score indicates an unhealthier FOPL MTL score

**Supplementary Table 9: Binary regression modelling the association between NOVA group and FOPL MTL score (median and below vs. above median cutoff)**

|  | **Exp(Beta)** | **95% Confidence Interval** | | **p-value** |
| --- | --- | --- | --- | --- |
|  |  | **Lower** | **Upper** |  |
| **FOPL score** |  |  |  |  |
| UPF | 6.034 | 5.032 | 7.236 | <0.001 |
| PF and PCI | 5.384 | 4.137 | 7.009 | <0.001 |

Green FOPL traffic light = 1, Amber FOPL traffic light = 2, Red FOPL traffic light = 3. Four green FOPL traffic lights = 4, four red FOPL traffic lights = 12.

Reference = MPF

Higher score indicates greater odds of having an unhealthier FOPL MTL score

Median cut-off = 6

**Supplementary Table 10: The most common MPFs and UPFs with no red FOPL traffic lights**

| **MPF (n=820)** | **n** | **%** | **UPF (n=855)** | **n** | **%** |
| --- | --- | --- | --- | --- | --- |
| Other vegetables including homemade dishes | 109 | 13.3 | Sandwiches | 65 | 7.6 |
| Other fruit not canned | 85 | 10.4 | High fibre breakfast cereals | 43 | 5.0 |
| Salad and other raw vegetables | 66 | 8.0 | Other milk | 38 | 4.5 |
| Other beef & veal including homemade recipe dishes | 55 | 6.7 | White bread (not high fibre; not multiseed bread) | 35 | 4.1 |
| Other chicken / turkey including homemade recipe dishes | 49 | 6.0 | Savoury sauces pickles gravies & condiments | 29 | 3.4 |
| Beans and pulses including ready meal & homemade dishes | 37 | 4.5 | Soft drinks not low calorie ready to drink still | 27 | 3.2 |
| Other cereals | 33 | 4.0 | Yogurt | 26 | 3.0 |
|  |  |  | Manufactured beef products including ready meals | 24 | 2.8 |
|  |  |  | Biscuits manufactured / retail | 24 | 2.8 |
|  |  |  | Pasta manufactured products & ready meals | 23 | 2.7 |
|  |  |  | Pizza | 22 | 2.6 |
|  |  |  | Manufactured chicken products including ready meals | 22 | 2.6 |
|  |  |  | Meat alternatives including ready meals & homemade dish | 22 | 2.6 |
|  |  |  | Manufactured coated chicken / turkey products | 20 | 2.3 |
|  |  |  | White fish coated or fried | 20 | 2.3 |

**Supplementary Table 11: Nutrient content per 100g of UPFs with or without red FOPL traffic lights**

| **Nutrient** | **UPF with no red FOPL traffic lights (n=855)** | | **UPF with one or more red FOPL traffic lights (n=795)** | | **p-value** |
| --- | --- | --- | --- | --- | --- |
|  | **Median** | **IQR** | **Median** | **IQR** |  |
| Fat (g/100g) | 3.9 | 1.3, 8.0 | 15.7 | 6.6, 23.5 | < 0.001 |
| Saturated fat (g/100g) | 1.0 | 0.3, 2.1 | 5.9 | 1.7, 9.8 | < 0.001 |
| Total sugar (g/100g) | 3.0 | 1.5, 5.5 | 16.4 | 2.1, 37.9 | < 0.001 |
| Salt (g/100g) | 0.54 | 0.13, 0.92 | 0.65 | 0.23, 1.45 | < 0.001 |
| Energy (kcal/g) | 1.52 | 0.77, 2.43 | 3.53 | 2.51, 4.43 | < 0.001 |
| Protein (g/100g) | 6.4 | 2.1, 10.7 | 5.5 | 3.1, 9.2 | 0.761 |
| Water (g/100g) | 65.2 | 45.9, 80.9 | 27.3 | 5.4, 52.5 | < 0.001 |
| Fibre (g/100g) | 1.5 | 0.5, 3.0 | 1.5 | 0.4, 2.8 | 0.435 |

**Supplementary Table 12: Nutrient and energy content of items with no red FOPL traffic lights by NOVA group.**

| **Nutrient** | **TOTAL (n=1846)** | | **MPF (n=820)** | | **PCI (n=9)** | | **PF (n=162)** | | **UPF (n=855)** | | **p-value** |
| --- | --- | --- | --- | --- | --- | --- | --- | --- | --- | --- | --- |
|  | **Median** | **IQR** | **Median** | **IQR** | **Median** | **IQR** | **Median** | **IQR** | **Median** | **IQR** |  |
| Fat (g/100g) | 2.2 | 0.4, 6.6 | 1.0 (a) | 0.3, 4.5 | 0.0 (b) | 0.0, 0.2 | 3.5 (a) | 0.0, 7.9 | 3.9 (c) | 1.3, 8.0 | < 0.001 |
| Saturated fat (g/100g) | 0.6 | 0.1, 1.8 | 0.2 (a) | 0.1, 1.1 | 0.0 (b) | 0.0, 0.0 | 1.0 (c) | 0.0, 2.9 | 1.0 (c) | 0.3, 2.1 | < 0.001 |
| Total sugar (g/100g) | 2.6 | 1.0, 5.1 | 2.2 (a) | 0.4, 4.6 | 0.0 (b) | 0.0, 0.0 | 2.6 (c) | 1.3, 8.4 | 3.0 (c) | 1.5, 5.5 | < 0.001 |
| Salt (g/100g) | 0.18 | 0.02, 0.63 | 0.05 (a) | 0.01, 0.19 | 0.08 (ab) | 0.01, 0.50 | 0.32 (b) | 0.02, 0.72 | 0.54 (c) | 0.13, 0.92 | < 0.001 |
| Energy (kcal/g) | 1.09 | 0.44, 1.97 | 0.75 (a) | 0.32, 1.34 | 0.53 (abc) | 0.01, 2.54 | 1.08 (b) | 0.59, 1.75 | 1.52 (c) | 0.77, 2.43 | < 0.001 |
| Protein (g/100g) | 4.6 | 1.3, 10.9 | 3.2 (ab) | 1.1, 11.3 | 0.3 (ab) | 0.2, 23.5 | 4.1 (b) | 0.6, 10.1 | 6.4 (a) | 2.1, 10.7 | < 0.001 |
| Water (g/100g) | 74.3 | 57.0, 86.4 | 79.9 (a) | 68.2, 89.2 | 8.7 (b) | 2.5, 41.5 | 75.2 (a) | 65.0, 85.7 | 65.2 (b) | 45.9, 80.9 | 0.006 |
| Fibre (g/100g) | 1.4 | 0.4, 2.7 | 1.5 (a) | 0.0, 2.7 | 26.0 (a) | 0.0, 61.3 | 1.1 (b) | 0.1, 1.6 | 1.5 (a) | 0.5, 3.0 | < 0.001 |

Unlike letters indicates significantly different p < 0.05

Pairwise comparisons conducted using Kruskal-Wallis ANOVA with Bonferroni correction for multiple comparisons.

**Supplementary Table 13: Binary regression modelling the association between NOVA group and nutrient content of items with no red FOPL traffic lights (median and below vs. above median for each nutrient)**

|  | **Exp(B)** | **95% Confidence Interval** | | **p-value** |
| --- | --- | --- | --- | --- |
|  |  | **Lower** | **Upper** |  |
| **Fat (g/100g)** |  |  |  |  |
| UPF | 3.606 | 2.948 | 4.411 | < 0.001 |
| PF and PCI | 1.872 | 1.342 | 2.611 | < 0.001 |
| **Saturated fat (g/100g)** |  |  |  |  |
| UPF | 3.239 | 2.652 | 3.957 | < 0.001 |
| PF and PCI | 1.941 | 1.392 | 2.707 | < 0.001 |
| **Total sugar (g/100g)** |  |  |  |  |
| UPF | 1.563 | 1.288 | 1.895 | < 0.001 |
| PF and PCI | 1.136 | 0.816 | 1.582 | 0.450 |
| **Salt (g/100g)** |  |  |  |  |
| UPF | 6.307 | 5.099 | 7.800 | < 0.001 |
| PF and PCI | 3.469 | 2.471 | 4.870 | < 0.001 |
| **Energy (kcal/g)** |  |  |  |  |
| UPF | 2.942 | 2.412 | 3.588 | < 0.001 |
| PF and PCI | 1.56 | 1.119 | 2.175 | 0.009 |
| **Protein (g/100g)** |  |  |  |  |
| UPF | 1.950 | 1.605 | 2.368 | < 0.001 |
| PF and PCI | 1.229 | 0.883 | 1.711 | 0.222 |
| **Water (g/100g)** |  |  |  |  |
| UPF | 0.330 | 0.270 | 0.402 | < 0.001 |
| PF and PCI | 0.578 | 0.414 | 0.805 | 0.001 |
| **Fibre (g/100g)** |  |  |  |  |
| UPF | 1.039 | 0.858 | 1.258 | 0.698 |
| PF and PCI | 0.418 | 0.294 | 0.595 | < 0.001 |

Reference = MPF

Higher score indicates greater odds of having above median nutrient content vs. median or below

**Supplementary Table 14: Nutrient content per 100g of UPFs without red FOPL traffic lights and two or more green FOPL traffic lights vs. all others**

| **Nutrient** | **UPF with no red and at least two green FOPL traffic lights (n=554)** | | **UPF with red and/or less than two green FOPL traffic lights (n=1,096)** | | **p-value** |
| --- | --- | --- | --- | --- | --- |
|  | **Median** | **IQR** | **Median** | **IQR** |  |
| Fat (g/100g) | 2 | 0.5, 4.3 | 12.3 | 5.8, 21.1 | < 0.001 |
| Saturated fat (g/100g) | 0.5 | 0.1, 1.0 | 4.0 | 1.7, 8.2 | < 0.001 |
| Total sugar (g/100g) | 2.9 | 1.3, 5.1 | 6.7 | 2.0, 29.1 | < 0.001 |
| Salt (g/100g) | 0.33 | 0.08, 0.80 | 0.74 | 0.27, 1.28 | < 0.001 |
| Energy (kcal/g) | 1.07 | 0.48, 2.21 | 2.91 | 2.07, 4.11 | < 0.001 |

**Supplementary Table 15: Nutrient content per 100g of items without red FOPL traffic lights and two or more green FOPL traffic lights vs. all others, by NOVA group**

| **Nutrient** | **TOTAL (n=1403)** | | **MPF (n=738)** | | **PCI (n=9)** | | **PF (n=102)** | | **UPF (n=554)** | | **p-value** |
| --- | --- | --- | --- | --- | --- | --- | --- | --- | --- | --- | --- |
|  | **Median** | **IQR** | **Median** | **IQR** | **Median** | **IQR** | **Median** | **IQR** | **Median** | **IQR** |  |
| Fat (g/100g) | 1.2 | 0.2, 3.5 | 0.7 (a) | 0.2, 2.9 | 0.0 (b) | 0.0, 0.2 | 0.1 (b) | 0.0, 2.1 | 2.0 (c) | 0.5, 4.3 | < 0.001 |
| Saturated fat (g/100g) | 0.23 | 0.04, 0.90 | 0.16 (a) | 0.05, 0.74 | 0.0 (b) | 0.0, 0.03 | 0.02 (b) | 0.00, 0.77 | 0.50 (c) | 0.10, 1.01 | < 0.001 |
| Total sugar (g/100g) | 2.5 | 0.8, 5.1 | 2.2 (a) | 0.4, 4.8 | 0.0 (b) | 0.0, 0.0 | 3.3 (c) | 1.5, 10..0 | 2.9 (c) | 1.3, 5.1 | < 0.001 |
| Salt (g/100g) | 0.10 | 0.01, 0.35 | 0.03 (a) | 0.01, 0.17 | 0.08 (abc) | 0.01, 0.5 | 0.04 (b) | 0.01, 0.30 | 0.33 (c) | 0.08, 0.80 | < 0.001 |
| Energy (kcal/g) | 0.81 | 0.33, 1.61 | 0.71 (a) | 0.29, 1.24 | 0.53 (ab) | 0.01, 2.54 | 0.74 (a) | 0.40, 1.02 | 1.08 (b) | 0.48, 2.21 | < 0.001 |

Unlike letters indicates significantly different p < 0.05

Pairwise comparisons conducted using Kruskal-Wallis ANOVA with Bonferroni correction for multiple comparisons.

**Supplementary Table 16: Number of items defined as being hyper-palatable based on fat, sodium, sugar and carbohydrate content.**

| **Hyper-palatable cluster** | **MPF** | **PCI** | **PF** | **UPF** | **p-value** | **Total** |  |
| --- | --- | --- | --- | --- | --- | --- | --- |
| **FSOD** |  |  |  |  |  | **n** | **%** |
| **Yes** | 36 | 3 | 99 | 504 |  | 642 | 24.1% |
| **No** | 867 | 43 | 156 | 957 | <0.001 | 2023 | 75.9% |
| **FS** |  |  |  |  |  |  |  |
| **Yes** | 78 | 0 | 25 | 318 |  | 421 | 15.8% |
| **No** | 825 | 46 | 230 | 1143 | <0.001 | 2244 | 84.2% |
| **CSOD** |  |  |  |  |  |  |  |
| **Yes** | 9 | 1 | 13 | 342 |  | 365 | 13.7% |
| **No** | 894 | 45 | 242 | 1119 | <0.001 | 2300 | 86.3% |
| **Overall (FSOD and/or FS and/or CSOD)** |  |  |  |  |  |  |  |
| **Yes** | 121 | 4 | 127 | 994 |  | 1246 | 46.8% |
| **No** | 782 | 42 | 128 | 467 | <0.001 | 1419 | 53.2% |

FSOD: (1) fat and sodium (> 25% kcal from fat, ≥ 0.30% sodium content by weight); FS: (2) fat and simple sugars (> 20% kcal from fat, > 20% kcal from sugar) CSOD (3) carbohydrates and sodium (> 40% kcal from carbohydrates, ≥ 0.20% sodium by weight); any of the three clusters.

**Supplementary Figure 1: The number of red or green FOPL traffic lights within MPFs or UPFs.**

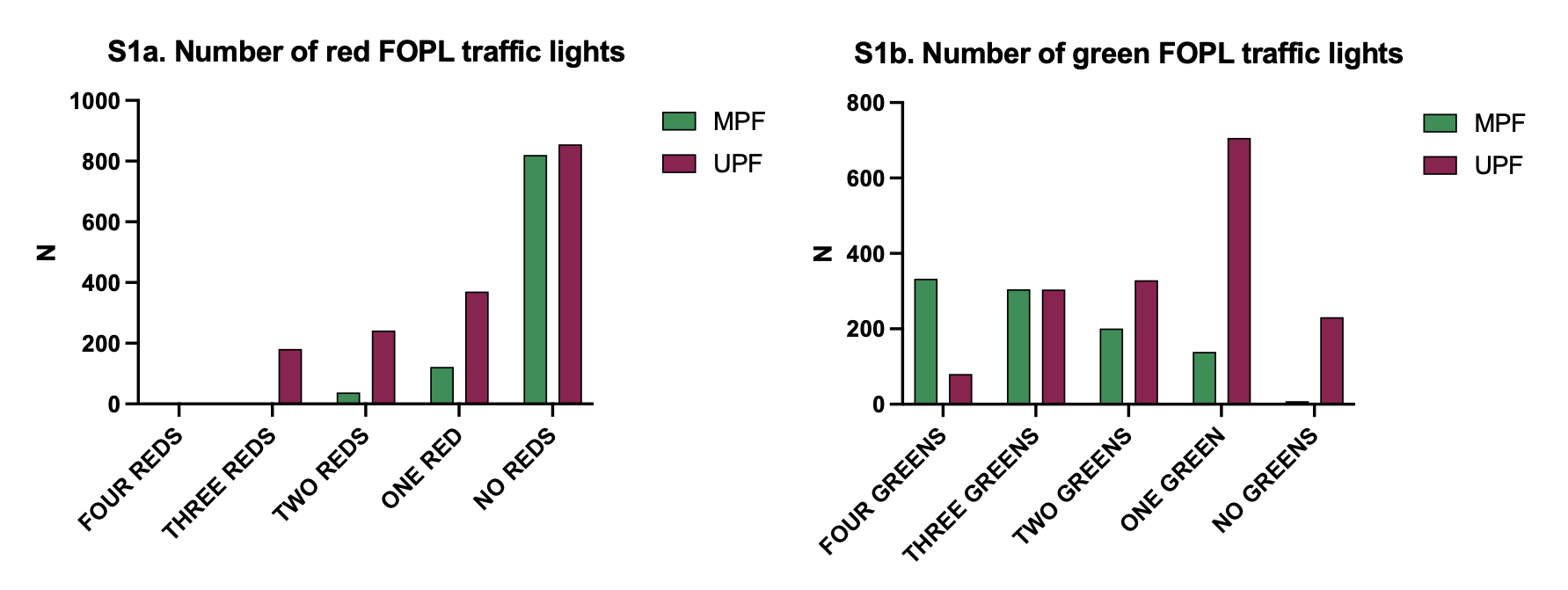

**Supplementary Figure 2: FOPL MTL score within MPFs or UPFs.**

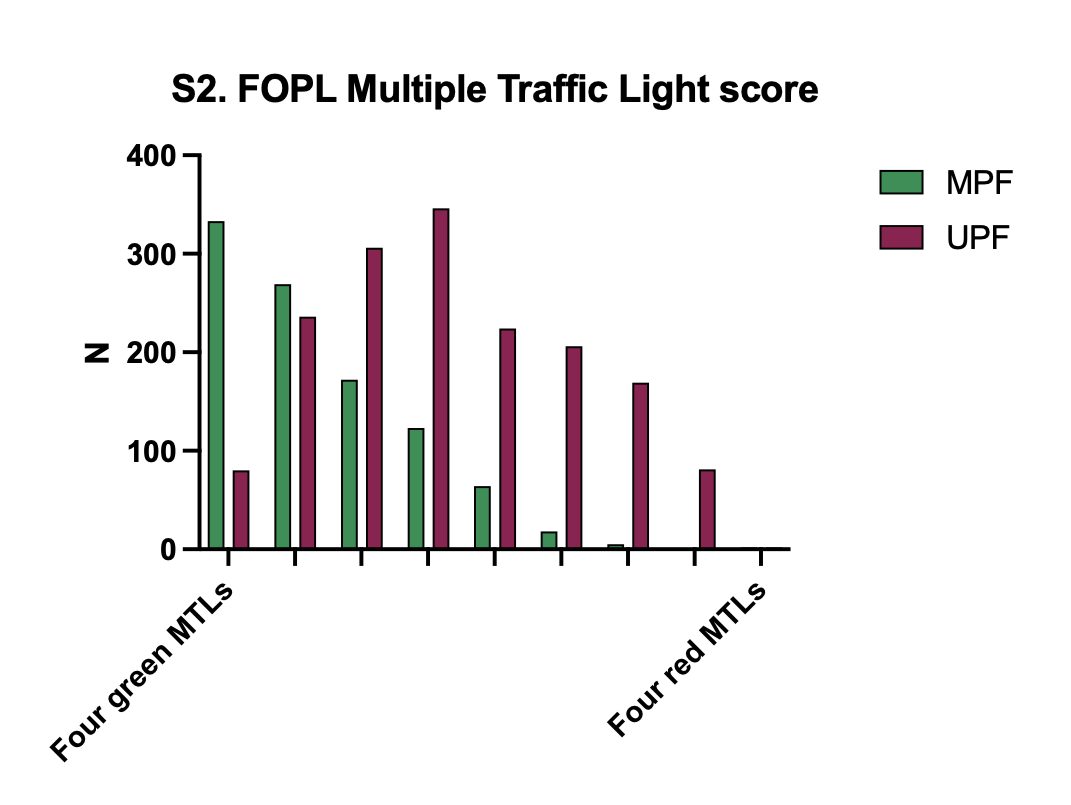
